# Cytokine and chemokine gene expression in patients with clinically suspect arthralgia; a longitudinal study during progression to inflammatory arthritis or non-progression

**DOI:** 10.1101/2022.08.26.22279253

**Authors:** J.W. Heutz, C. Rogier, E. Niemantsverdriet, S.J.F. van den Eeden, P.H.P. de Jong, E. Lubberts, A. Geluk, A.H.M. van der Helm-van Mil

**Affiliations:** Department of Rheumatology, Erasmus Medical Center Rotterdam, The Netherlands; Department of Rheumatology, Leiden University Medical Center, The Netherlands

## Abstract

**Background:** Autoantibody-responses rise years before onset of inflammatory arthritis (IA) and are stable during transitioning from clinically suspect arthralgia (CSA) to IA. Cytokine and chemokine levels can also rise years before IA-onset, but the course after CSA-onset is unknown. To better understand the processes in this symptomatic at-risk phase, we studied the course of cytokine, chemokine and related receptors gene expression in CSA-patients during progression to IA, and in CSA-patients who ultimately did not develop IA. Differential expressed genes between ACPA-positive and ACPA-negative CSA-patients who developed IA were also explored.

**Methods:** Whole blood RNA expression of 37 inflammatory cytokines/chemokines/related receptors was determined by dual-color reverse-transcription multiplex ligation-dependent probe amplification, in paired samples of CSA-patients at CSA-onset and either at IA-development or after 24-months without IA-development. ACPA-positive and ACPA-negative CSA-patients developing IA were compared at CSA-onset and during progression to IA. GEE-models tested changes over time. A false discovery rate approach was applied to correct for multiple testing.

**Results:** None of the cytokines/chemokine genes significantly changed in expression between CSA-onset and IA-development. In CSA-patients without IA development, G-CSF expression decreased (p=0.001), whereas CCR6 and TNIP expression increased (p<0.001 and p=0.002, respectively) over a 2-year period. Expression levels in ACPA-positive and ACPA-negative CSA-patients who developed IA were similar.

**Conclusion:** Whole blood gene expression of cytokines, chemokines and related receptors did not change significantly from CSA to IA-development. This suggests that changes in expression of these molecules occurred preceding CSA-onset and may not relate to the final hit of developing chronic arthritis. Observed changes in CSA-patients not developing IA can provide clues for processes related to resolution.

## Introduction

Rheumatoid arthritis (RA) is a chronic autoimmune disease, that develops gradually [1]. In the majority of patients, the onset of clinically apparent inflammatory arthritis (IA) is preceded by a phase of arthralgia. A combination of signs and symptoms that is suspect for progression to RA can be recognized by rheumatologists and is called clinical suspect arthralgia (CSA) [2]. Approximately twenty percent of patients with CSA will progress to IA; the period between symptom onset and IA-development generally takes 6-12 months [3]. To date, the final hit or final process in the pathophysiology related to progression from CSA to IA is still unclear.

In the pathophysiology of disease development, autoantibody-responses occur and rise years before the onset of IA. Their levels are stable during the transition from clinically suspect arthralgia (CSA) to IA. In addition, the levels are stable in autoantibody-positive CSA-patients who over time do not develop IA [4-6]. Together, this suggests that auto-antibody maturation occurs before the joint symptoms occur and does not relate to the final hit of IA development.

In contrast to the autoantibody response, which has extensively been studied in the phases preceding IA, less is known about the course of inflammatory proteins such as cytokines and chemokines in this phase. Some nested case-control studies found upregulation of different inflammatory markers in the pre-IA phase compared with healthy controls [7-9]. One nested case-control study included blood samples from 85 patients preceding IA onset; this showed upregulation of several cytokines and chemokines compared to control subjects, among which were IFNγ, IL-13, CXCL9, CCL11 and IL-1Ra [10]. Sokolove *et al* reported in a longitudinal study the rise and stabilization of multiple cytokine and chemokine levels years before IA-onset [11]. However, the above mentioned studies did not show whether the changes in cytokines expression occurred already in the asymptomatic pre-disease stage or occurred after symptom development, thus during progression from CSA to clinical arthritis. The relationship with symptom onset is relevant because symptoms can be recognized in a clinical setting where intervention is possible. Ideally intervention in this stage is targeted at processes that are fundamental for progression to chronic disease. In addition, none of the nested case-control studies included a control group in a similar at-risk stage that ultimately did not develop IA. A control group in a similar at-risk stage is relevant to identify processes that are specific for progression from arthralgia to chronic disease.

In this longitudinal study in CSA-patients, we addressed three questions with the ultimate aim to increase the understanding of the processes in the symptomatic at-risk phase of CSA. First, we studied the course of cytokine, chemokine and related receptor gene expression in CSA during progression to IA. Based on the results from above mentioned studies on autoantibody responses [4-6, 11], we hypothesized that also rises in cytokine and chemokine expression mostly precede the onset of CSA. Second, this study is the first to determine the course of cytokine and chemokine and related receptor gene expression in CSA-patients that did not develop IA. Finally, we explored whether there were differences between ACPA-positive and ACPA-negative CSA patients that progressed to IA, at CSA-onset and over time. This was done since studies on risk-factors suggest that autoantibody positive and autoantibody negative RA are subgroups with differences in the underlying pathophysiology [12].

## Methods

### Study population

Patients were obtained from the Leiden CSA cohort [2]. In the Leiden CSA cohort, all patients that presented at the outpatient clinical with recent onset (<6 months) arthralgia of the small joints, suspicious for progression to RA according to a rheumatologist, were consecutively included. Baseline visits consisted of physical examination, blood sampling (including PAX gene tubes) and imaging of hands and feet. Autoantibody status was not known at time of inclusion, since general practitioners were discouraged to test for them according to Dutch guidelines. Follow up research visits took place at 4, 12 and 24 months. In case of joint swelling or gain of symptoms, additional visits were scheduled. Patients were followed for two years or until IA development, determined by a rheumatologist at physical examination (66 swollen joint count ≥ 1). Treatment with disease modifying anti-rheumatic drugs (DMARDs), including corticosteroids, was not allowed during follow up.

A detailed flowchart of the included patient samples is depicted in Figure 1. In this study, RNA expression of 37 cytokines and chemokines was measured using blood from PAX gene tubes of CSA patients. Between April 2012 and January 2019 639 patients were included in the Leiden CSA cohort. PAX gene tubes were not available from 44 patients at baseline. 98 patients were included in a randomized placebo-controlled trial and were excluded, because of possible DMARD treatment. RNA measurements failed for 2 patients. In total, from 495 eligible patients baseline RNA measurements were obtained.

**Figure 1.**
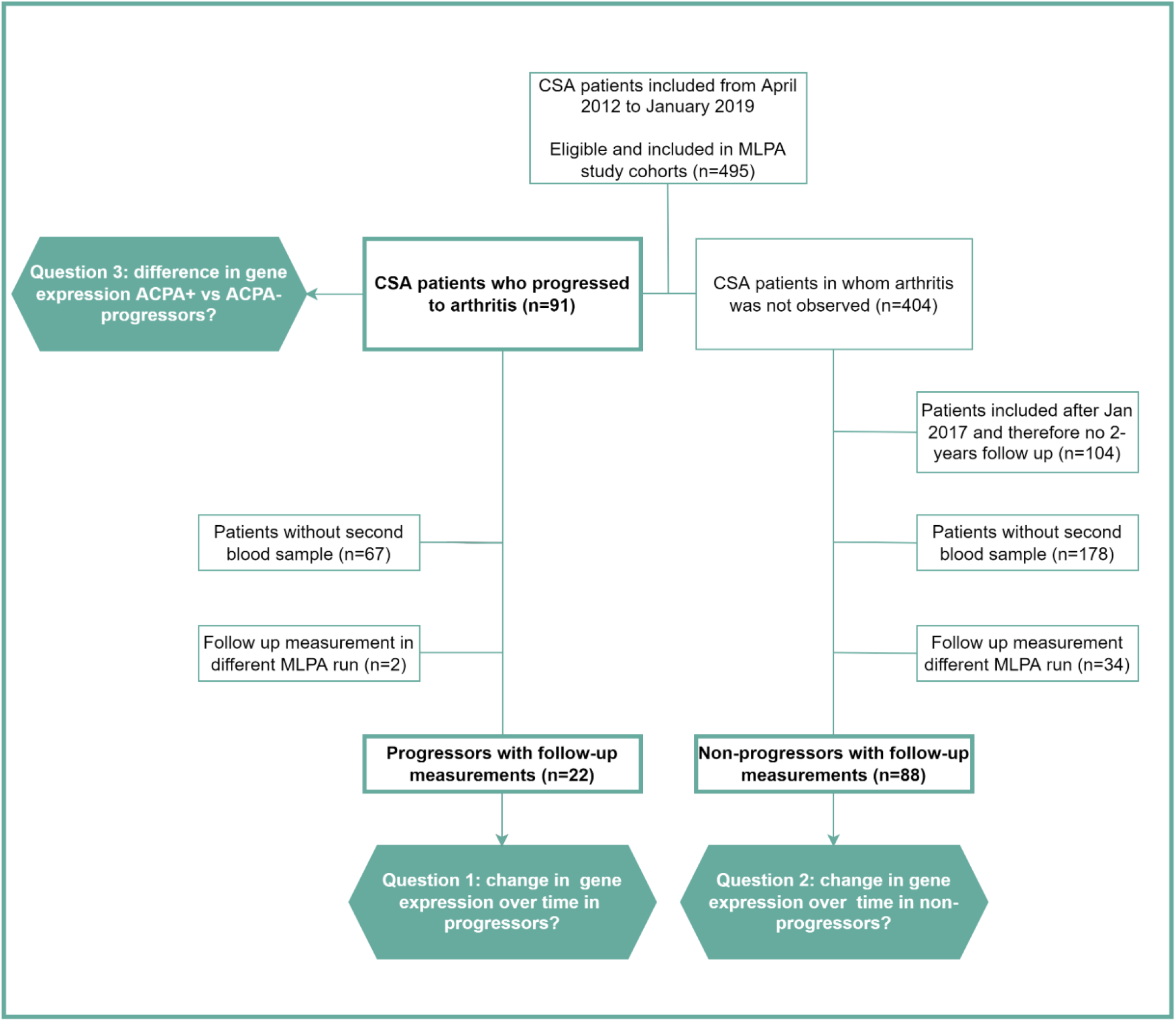
Flowchart of included patients in the three analyses belonging to the three study questions

From 22 out of 91 CSA patients that progressed to arthritis PAX gene tubes were available at both baseline and at time of arthritis development to detect changes in expression over time (n=67 without a second PAX gene tube, n=2 with paired MLPA measurements in different runs and therefore excluded from longitudinal analyses). From 404 patients that did not progress to arthritis, all patients included after January 2017 were excluded because of lack of 2 years follow up (n=104). From 88 out of 404 patients that did not progress to arthritis PAX gene tubes were available at both baseline and after 2 years to detect changes in gene expression over time. Absence of a second PAX gene tube was mostly due to logistic factors (n=178). Patients with paired MPLA measurements in different runs were excluded from longitudinal analyses (N=34). Baseline characteristics of patients with and without a paired sample are provided in Supplemental Table 1. Except for a small difference in ACPA-positivity between progressing CSA-patients with and without a second sample (27% versus 44% respectively), baseline characteristics did not remarkably differ.

### Measurement of RNA expression levels

#### RNA isolation

From baseline and follow up, whole blood in PAXgene tubes RNA was extracted using PAXgene Blood RNA kits (BD Biosciences, Franklin Lakes, N) according to the manufacturers’ protocol [13]. RNA yield was measured by NanoDrop ND-1000 spectrophotometer (NanoDrop Technologies, Wilmington, DE).

#### Dual color reverse-transcription multiplex ligation-dependent probe amplification (dcRT-MLPA) assays

RNA expression was determined by dcRT-MLPA as previously described [14]. RNA expression analysis included 37 cytokines, chemokines and related receptors: CXCR5, CCL11, CCL17, CCL18, CCL2, CCL22, CCL3, CCL4, CCR6, CCR7, CSF2, CSF3, CXCL10, CXCL3, CXCL4, CXCL7, CXCL9, IFNG, IL10, IL12A, IL12B, IL13, IL15, IL18, IL1A, IL1B, IL1Ra, IL2, IL22RA1, IL23A, IL6, IL7R, IL8, IL9, TGFB, TNF, TNIP1. Data were analyzed using GeneMapper software 5 (Applied Biosystems). Results from target genes were calculated relative to the geometric average signal of housekeeping gene glyceraldehyde 3-phosphate dehydrogenase (GAPDH) [15]. Signals below the noise cutoff (log2 transformed peak area ≤7.64) were assigned the threshold value. Further analyses were performed on log2 transformed data.

RNA measurements on baseline blood samples were performed in 2 runs. The first run comprised blood samples from patients included between April 2012 and March 2015 (n=236). The second run comprised blood samples from patients included between March 2015 and January 2019 (n=259). Follow up measurements (either at time of arthritis development or after 2 years) were performed in either MLPA run 1 or 2.

### Sub-study with healthy controls

Although the group of CSA-patients that did not progress to IA was the main reference group in this study, we performed a small sub-study in which we determined the expression of genes known to be differently expressed in the pre-RA phase (IFNγ, IL-13, CXCL9, CCL11 and IL1Ra) compared to healthy controls. These genes were determined in a (separate) MLPA run in CSA patients that progressed to arthritis (progressors, n=4), CSA patients that did not progress to arthritis (non-progressors, n=6) and healthy controls (n=5). This was done to validate our assay with results known from previous studies [10].

### Statistical analyses

Generalized estimated equations were used to test average changes in cytokine and chemokines expression levels over time within patients that progressed to IA and, separately, in patients that did not progress to IA. In patients that progressed to IA, a sensitivity analysis of gene expression over time was conducted in patients who were diagnosed with RA (i.e. with a clinical diagnosis and who fulfilled the 1987 and/or 2010 criteria for RA and/or started a DMARD at IA-development; n=18). Logistic regression models were used to test baseline differences in gene expression levels between ACPA positive (n=40) and ACPA negative (n=51) patients that progressed to IA. All analyses were corrected for the two different MLPA runs. No other corrections were made. False discovery rate (FDR) was used to correct for multiple testing. P-values ≤0.05 were considered statistically significant. STATA version 17.0 was used.

## Results

### Study population

Baseline characteristics of the patients who were studied for the three research questions are presented in Table 1.

**Table 1.** Baseline characteristics of CSA-patients included in three analyses; (1) CSA-patients that progressed to IA with paired measurements; (2) CSA-patients who did not progress with paired measurements over 2 years and; (3) CSA-patients who progressed to IA stratified for ACPA status

|  | Progressors<br>with paired<br>measurements | Non-progressors<br>with paired<br>measurements | Comparison of ACPA+ and<br>ACPA- progressors |  |
| --- | --- | --- | --- | --- |
|  | n=22 | n=88 | ACPA+ n=40 | ACPA- n=51 |
| Age in years, mean (SD) | 49 (12) | 44 (12) | 48 (13) | 46 (13) |
| Gender, female, n (%) | 17 (77) | 71 (81) | 32 (80) | 35 (31) |
| Symptom duration in<br>weeks, median (IQR) | 19 (7-53) | 23 (14-46) | 29 (15-54) | 15 (7-34) |
| Tender joint count,<br>median (IQR) | 5 (3-7) | 5 (2-10) | 4 (2-7) | 5 (4-10) |
| ACPA positivity, n (%) | 6 (27) | 9 (10) | 40 (100) | 0 (0) |
| RF positivity, n (%) | 7 (32) | 14 (16) | 36 (90) | 9 (18) |
| Increased CRP, n (%) | 7 (32) | 10 (11) | 13 (33) | 17 (33) |
| MRI-detected subclinical<br>inflammation, n (%) | 14 (64) | 31 (36) | 32 (89) | 34 (68) |

CSA, clinical suspect arthralgia; IA, inflammatory arthritis; ACPA, anti-citrullinated protein antibody; RF, rheumatoid factor; MRI, magnetic resonance imaging; CRP, c-reactive protein

### Sub-study with known differently expressed genes to healthy controls

Figure 2 gives a visual representation of genes in CSA-patients that progressed to IA and healthy controls. In line with earlier studies in pre-IA [10, 11], CSA patients showed increased expression of IFNɣ, IL-13, CXCL9, CCL11 and IL-1Ra compared to healthy controls. In addition, our data also show that CSA-patients that did not progress to IA also showed higher expression of these genes compared to healthy controls.

**Figure 2.**
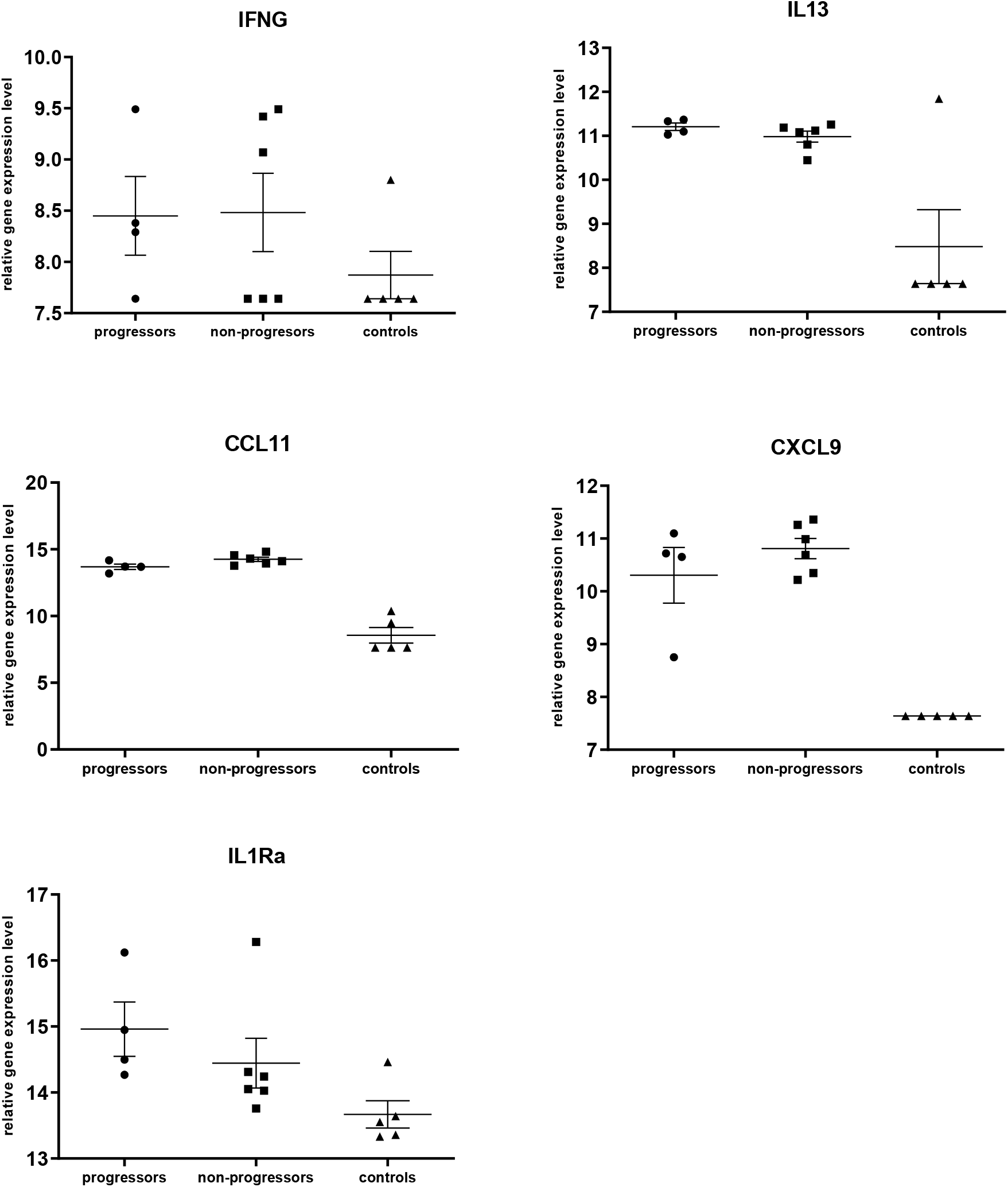
Gene expression of IFNG, IL13, CCL11, CXCL9, and IL1Ra in CSA-patients that did progress to IA vs CSA patients that did not progress to RA vs healthy controls.

### Cytokine, chemokine and receptor gene expression over time from CSA-onset to IA-development

The median time between presentation with CSA and IA-development was 4.1 months (IQR 3.4-8.3). After correction for multiple testing and MLPA run, none of the 37 cytokine and chemokine gene expression levels significantly changed over time towards IA-development. The course over time is depicted in Figure 3; p-values are presented in Supplemental Table 2. Sensitivity analysis in patients that progressed to RA showed similar results (Supplemental Figure 1).

**Figure 3.**
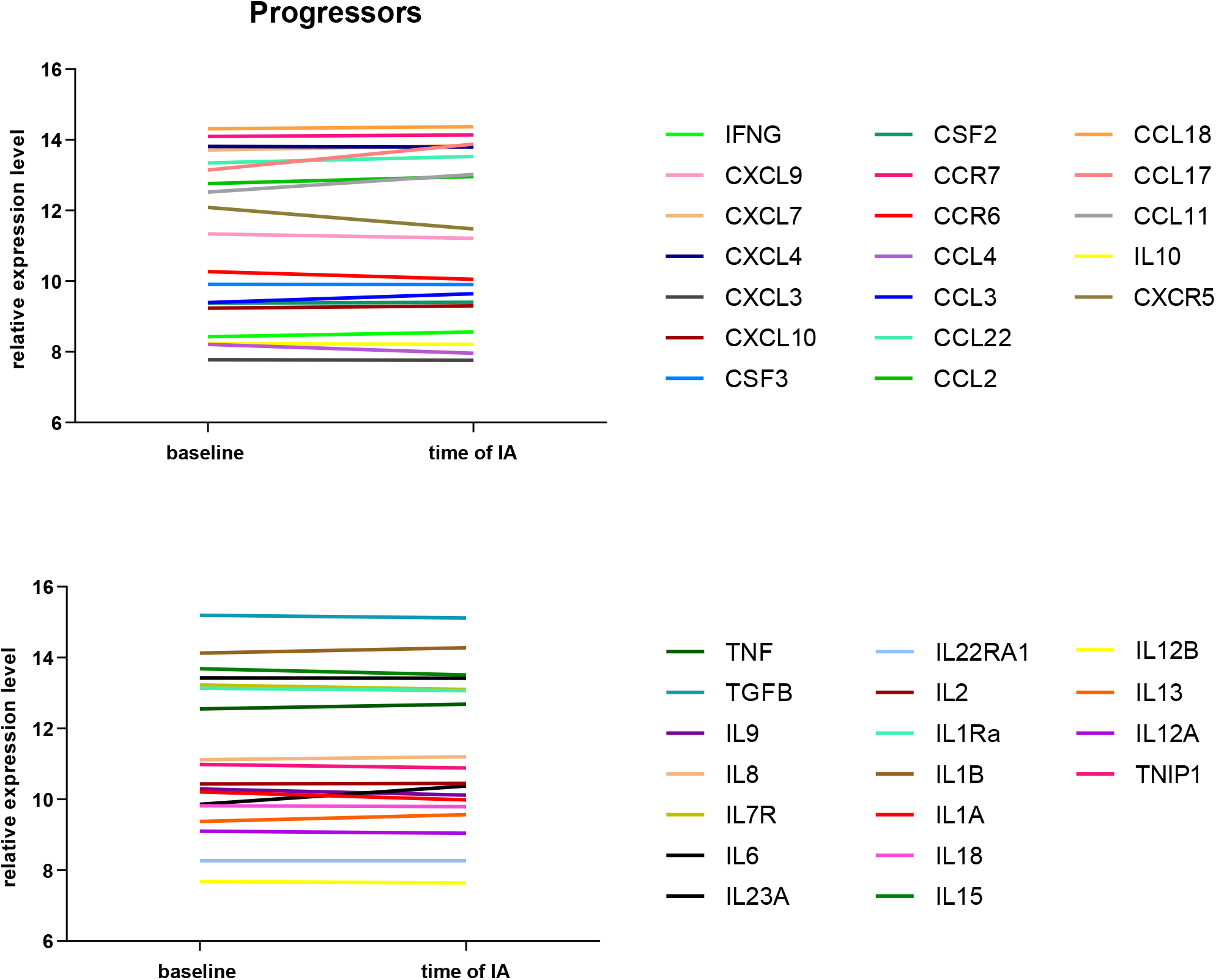
Modelled course of gene expression of 37 cytokines, chemokines and related receptors in CSA patients that progressed to IA. Cytokines and chemokines were measured at baseline and at time of IA development and for reasons of clarity presented in two plots. No statistically significant changes were observed during follow-up. CSA, clinical suspect arthralgia; IA, inflammatory arthritis.

**Figure 4.**
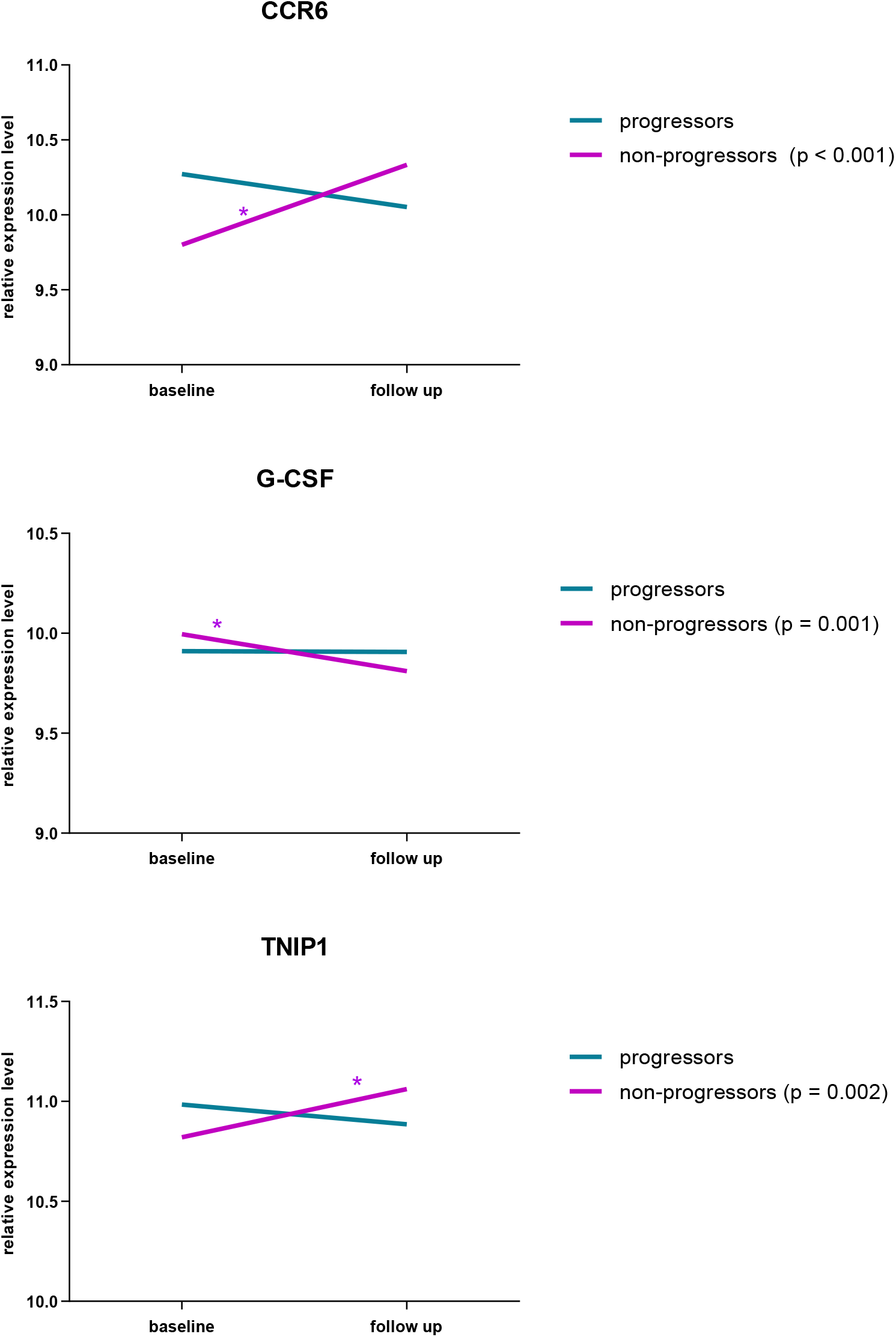
Modelled course of gene expression of G-CSF, CCR6 and TNIP1 in patients that did not progress to arthritis with paired samples with 2-year intervals. For comparison the course of patients that progressed to IA was included, here the 2^nd^ samples was collected at IA development. IA, inflammatory arthritis.

### Cytokine, chemokine and receptor gene expression over time in CSA-patients that did not progress to IA

Patients that presented with CSA and over time did not progress to IA were studied with paired samples with a 2 year interval. The expression level of granulocyte colony-stimulating factor (G-CSF) had decreased after 2 years of follow up (*p* < 0.001). The expression levels of C-C motif chemokine receptor 6 (CCR6) and TNFAIP3 interacting protein 1 (TNIP1) had both increased after 2 years of follow up (*p* < 0.001 and *p* = 0.002, respectively; Figure 3 & Supplemental Table 2). The other 34 genes did not change significantly.

### Comparison of ACPA-positive and ACPA-negative CSA-patients that progressed to IA

Here we compared differences between the development of ACPA-positive and ACPA-negative IA. At time of CSA presentation, expression of the 37 candidate genes did not differ between ACPA-positive and ACPA–negative CSA patients. IL-13 and CCR7 gene expression levels at baseline were lower in ACPA-positive compared to ACPA-negative patients, but after correction for multiple testing these results were not significant (Supplemental Table 3). The course of gene expression of cytokines and chemokines over time during progression to IA stratified for ACPA status is shown in Supplemental Figure 2. Visual inspection revealed no obvious differences and because of the low number of patients in the ACPA-positive group, no statistical tests were performed.

## Discussion

In this longitudinal study we aimed to increase the understanding of the processes involved in progression from the CSA at-risk stage to clinically apparent IA by studying the course of expression of cytokines, chemokines and related receptor genes, and relate this to the course of these molecules in CSA-patients that did not progress to IA. First, we observed that gene expression levels of cytokines and chemokines and related receptors did not change over time from CSA to IA-development. Second, in CSA-patients not developing IA, G-CSF, CCR6, and TNIP1 changed over the course of two years. Finally, gene expression at CSA onset and before development of clinical arthritis did not differ between ACPA-positive and ACPA-negative disease.

Our main finding that gene expression levels of 37 cytokines, chemokines and receptors did not change during the transition from arthralgia to IA, suggests that changes in these genes/molecules studied did not relate to the final hit of developing IA. In combination with the result from our sub-study, which suggested that gene expression levels from IFNγ, IL-13, CXCL9, CCL11 and IL-1Ra are lower in healthy controls compared to CSA-patients that developed IA, the results imply that rises in cytokine and chemokine levels precede the onset of CSA. The observation that gene expression levels did not change towards IA-development is in line with previous literature [7-10, 16]. Although information on the presence/absence of symptoms in the pre-RA stage was not included, the results from previous nested-cased control studies suggest that rises in cytokine and chemokine levels occurred several years before the diagnosis of RA [7-10, 16]. Since CSA usually develops ∼6months before clinical arthritis onset, it is presumable that measured rises in cytokines/chemokines most likely took place before symptom development.

Interestingly, the current data on cytokines/chemokines also resemble the data on autoantibody maturation as measured in a longitudinal study in CSA, that showed that measured autoantibody levels and number of isotypes and fine specificities were already increased at CSA-onset and did not mature any further during progression to IA [4]. In the absence of observed evident changes in markers from the systemic circulation during progression from CSA to IA, it can be hypothesized that the final decisive hit in the process of IA-development occurs locally, e.g. in the joints, in the phase of CSA. However, this is a subject for future studies. In parallel to arthritis development, leprosy is another chronic disease of which the development know several phases. In a longitudinal study in leprosy, the expression of genes encoding several inflammatory proteins increased preceding the onset of symptoms [17]. The order of a systemic immune response before abnormalities in the target organ occur may therefore be a shared trajectory between diseases.

To our knowledge, we are the first to study systemic expression of cytokines and chemokines in a symptomatic at-risk population that ultimately does not develop IA. In CSA patients not developing IA, gene expression of G-CSF had decreased and expression of CCR6 and TNIP1 had increased after two years. Although all the other genes did not change over the course of 2 years, it is likely that some of these genes were increased in expression at CSA presentation, since the CSA patients in our sub-study not progressing to clinical arthritis had higher levels of gene expression compared to healthy controls. The identification of G-CSF, CCR6, and TNIP1 that did change over the course of 2 years in our data can provide clues for processes mediating spontaneous resolution of CSA and related (subclinical) inflammation.

G-CSF expression levels in our study had decreased after 2 years. G-CSF is a pro-inflammatory cytokine involved in hematopoiesis that plays a role in inflammatory diseases, including RA [18]. Elevated levels of G-CSF have been detected in synovial fluid and serum of RA patients and are related to disease severity [19]. In a mouse model of RA, G-CSF levels increased over the course of the disease, blocking G-CSF ameliorated disease in mice. G-CSF deficiency protected mice from acute and chronic arthritis [20, 21]. This evidently pro-inflammatory role of G-CSF in established RA is in line with our finding that G-CSF decreased towards non-progression in CSA. Kokonnen *et al* reported in a nested case control study an elevated level of G-CSF in pre-RA patients compared to control subjects that did not rise further after diagnosis [10]. They did not include symptomatic at-risk patients that did not progress to arthritis, but this is in line with our finding that G-CSF did not change over time in CSA-patients that did progress to arthritis.

Gene expression levels of CCR6 had increased after 2 years in CSA-patients who did not progress to IA. A role for CCR6 in RA-development has been suggested from genetic studies where single nucleotide polymorphisms of CCR6 have been associated with rheumatoid arthritis development [22, 23]. Furthermore, increased proportions of peripheral blood CCR6+ Th17 cells have been reported in early RA and the CCR6/CCL20 axis plays an important role in the disease and the migration of Th17 cells to the joints [24, 25]. To our knowledge, no studies to date have investigated the role of this axis in the CSA phase. There is a possibility that migration of pro-inflammatory CCR6+ cells to the joint occurs before CSA-onset and that the rise in CCR6 expression in patients that do not progress to arthritis in our data represents resolution of this process in those patients. Another explanation could be that expression of regulatory/anti-inflammatory CCR6+ T-cells accounted for the rise in CCR6+ expression in CSA patients that did not progress to IA.

The third gene for which increased expression was detected after 2 years in CSA patients who did not progress to IA was TNIP1. TNIP1 encodes the protein ABIN1, which inhibits NF-kB signaling and plays a role in normal tissue homeostasis to prevent autoimmunity and inflammation [26]. Polymorphisms of TNIP1 have been associated with RA, as well as other polymorphisms of genes involved in nuclear factor-kB (NF-kB) signaling [27]. The increased expression of TNIP1 over time in our CSA-patients who did not progress to IA is in line with its anti-inflammatory role in the prevention of autoimmune diseases.

In our comparison of ACPA-positive and ACPA-negative CSA-patients transitioning to clinical arthritis, cytokine, chemokines and receptor expression levels did not differ at presentation with CSA. The course was evaluated visually, and appeared to reveal no differences over time, although no statistical inferences were made, because of the small number of patients in each group. Most previous studies in pre-RA patients on cytokines included mostly autoantibody-positive patients, and did not make direct comparisons to developing autoantibody-negative disease for individual cytokines [8, 10]. If validated in future studies, the similarity in cytokine and chemokines expression levels in ACPA-positive and ACPA-negative patients is interesting, since ACPA-positive and ACPA-negative RA have differences in environmental and genetic risk factors, but a largely similar clinical presentation at CSA onset and at RA diagnosis [12, 28, 29]. This could be suggestive of a common pathway in clinical arthritis development, despite differences in risk factors and in the severity of the disease after RA diagnosis.

Limitations of our study are the limited number of included patients in the longitudinal analysis of CSA patients that progressed to IA, and the absence of healthy control subjects in our main analysis. The limited number of patients may have led to false negative results regarding the detection of cytokines and chemokines that changed over time because of a lack of power. In addition, we studied a relatively small number of inflammatory genes and might therefore have missed other inflammatory genes that perhaps do change during progression from CSA to IA or RA. Another limitation is the fact that without healthy control subjects in our main analysis, we were not able to test for differences between CSA patients and healthy subjects. However, in a small sub-study we were able to compare progressing and non-progressing CSA patients to healthy controls. We demonstrated that observed changes in expression from the literature were also present in the current data, which shows validity of the findings. Moreover, our control group consisted of CSA-patients with similar symptoms but who did not pass the final step in transitioning to IA. In our view this group is valuable to understand the processes related to the final hit of IA development in already symptomatic patients. Finally, a limitation of our study is that the expression levels were measured at RNA level. Expression at RNA level does not necessarily translate to protein expression, which is more directly related to the actual activity. However, previous literature reporting increased protein levels of G-CSF in RA patients does not contradict our finding that CSA patients who do not develop IA show a decline in G-CSF RNA expression.

## Conclusions

In the transition from CSA to IA or RA, no changes in cytokine, chemokine or related receptor gene expression were observed in our study. Hence, the results from this study are the first to indicate that changes in systemic expression of these cytokines and chemokines are not related to the final hit of developing chronic RA and may therefore precede the onset of symptoms. Moreover, changes over time in at-risk patients not developing IA provided clues to processes related to spontaneous resolution of CSA/inflammation, which should be validated and expanded in future studies.

## Data Availability

All data produced in the present study are available upon reasonable request to the authors

## Ethics approval and consent to participate

The study was approved by the local medical ethical committee Leiden-Den Haag-Delft (P11.210). All patients gave written informed consent.

