## Supplemental Material for "Cytokine and chemokine gene expression in patients with clinically suspect arthralgia; a longitudinal study during progression to inflammatory arthritis or non-progression"

**Supplemental Table 1.** Baseline characteristics of (1) CSA patients who progressed to IA with and without paired measurements and (2) CSA patients who did not progress to IA with and without paired measurements

| | Progressors | | Non-progressors | |
|---|---|---|---|---|
| | With measurements over time n=22 | Without measurements over time n=91 | With measurements over time n=88 | Without measurements over time n=404 |
| Age in years, mean (SD) | 49 (12) | 47 (13) | 44 (12) | 43 (12) |
| Gender, female, n (%) | 17 (77) | 67 (74) | 71 (81) | 321 (79) |
| Symptom duration in weeks, median(IQR) | 19 (7-53) | 20 (9-52) | 23 (14-46) | 22 (10-52) |
| Tender joint count, median (IQR) | 5 (3-7) | 5 (2-8) | 5 (2-10) | 5 (2-10) |
| ACPA positivity, n (%) | 6 (27) | 40 (44) | 9 (10) | 24 (6) |
| RF positivity, n (%) | 7 (32) | 45 (49) | 14 (16) | 53 (13) |
| Increased CRP, n (%) | 7 (32) | 30 (33) | 10 (11) | 69 (17) |
| MRI-detected subclinical inflammation, n (%) | 14 (64) | 66 (77) | 31 (36) | 131 (34) |

Abbreviations: CSA, clinical suspect arthralgia; ACPA, anticitrullinated protein antibody; RF, rheumatoid factor; MRI, magnetic resonance imaging: CRP, c-reactive protein

| **Supplemental Table 2.** Results of GEE models evaluating the course of gene expression over time, in CSA patients who progressed to IA and who did not progress separately. | | | | | | |
|---|---|---|---|---|---|---|
| | **Progressors** | | | **Non-progressors** | | |
| **Gene** | **beta coefficient** | **p-value** | **FDR adjusted p-value** | **beta coefficient** | **p-value** | **FDR adjusted p-value** |
| BLR1 | -0.62 | 0.21 | 0.71 | -0.05 | 0.68 | 0.86 |
| CCL11 | 0.50 | 0.01 | 0.18 | 0.12 | 0.50 | 0.74 |
| CCL17 | 0.73 | 0.53 | 0.92 | -0.52 | 0.38 | 0.66 |
| CCL18 | 0.06 | 0.75 | 0.96 | 0.03 | 0.90 | 0.98 |
| CCL2 | 0.20 | 0.24 | 0.71 | -0.04 | 0.87 | 0.98 |
| CCL22 | 0.18 | 0.05 | 0.34 | 0.10 | 0.02 | 0.18 |
| CCL3 | 0.24 | 0.04 | 0.34 | 0.01 | 0.94 | 0.98 |
| CCL4 | -0.25 | 0.21 | 0.71 | 0.06 | 0.35 | 0.64 |
| CCR6 | -0.22 | 0.24 | 0.71 | 0.53 | <0.001 | <0.001 |
| CCR7 | 0.04 | 0.66 | 0.93 | -0.02 | 0.67 | 0.86 |
| CSF2 | 0.03 | 0.76 | 0.96 | -0.06 | 0.32 | 0.63 |
| CSF3 | 0.00 | 0.96 | 0.96 | -0.18 | <0.00 | 0.001 |
| CXCL10 | 0.07 | 0.59 | 0.93 | 0.11 | 0.42 | 0.67 |
| CXCL3L1 | -0.02 | 0.91 | 0.96 | -0.04 | 0.65 | 0.86 |
| CXCL4 | -0.02 | 0.89 | 0.96 | 0.06 | 0.33 | 0.63 |
| CXCL7 | 0.13 | 0.46 | 0.92 | 0.12 | 0.10 | 0.33 |
| CXCL9 | -0.13 | 0.54 | 0.92 | -0.22 | 0.16 | 0.44 |
| IFNG | 0.13 | 0.05 | 0.34 | 0.17 | 0.29 | 0.63 |
| IL10 | -0.03 | 0.92 | 0.96 | 0.01 | 0.93 | 0.98 |
| IL12A | -0.05 | 0.56 | 0.92 | 0.09 | 0.27 | 0.63 |
| IL12B | -0.03 | 0.32 | 0.80 | 0.00 | 0.98 | 0.98 |
| IL13_5RA | 0.19 | 0.47 | 0.92 | -0.18 | 0.06 | 0.30 |
| IL13_IFN | -0.12 | 0.01 | 0.18 | 0.03 | 0.51 | 0.74 |
| IL15 | -0.17 | 0.66 | 0.93 | 0.11 | 0.10 | 0.33 |
| IL18 | -0.02 | 0.95 | 0.96 | 0.48 | 0.05 | 0.30 |

| | | | | | | |
|---|---|---|---|---|---|---|
| IL1A | -0.22 | 0.64 | 0.93 | -0.40 | 0.09 | 0.33 |
| IL1B | 0.15 | 0.09 | 0.46 | 0.03 | 0.56 | 0.79 |
| IL1Ra | -0.07 | 0.53 | 0.92 | -0.04 | 0.71 | 0.87 |
| IL2 | 0.02 | 0.91 | 0.96 | -0.16 | 0.05 | 0.30 |
| IL22RA1 | 0.00 | 0.92 | 0.96 | 0.02 | 0.74 | 0.88 |
| IL23A | -0.01 | 0.83 | 0.96 | 0.00 | 0.97 | 0.98 |
| IL6_5RA | 0.52 | 0.01 | 0.18 | 0.31 | 0.07 | 0.30 |
| IL7R | -0.13 | 0.41 | 0.91 | 0.13 | 0.07 | 0.30 |
| IL8 | 0.08 | 0.71 | 0.96 | -0.08 | 0.41 | 0.67 |
| IL9 | -0.17 | 0.30 | 0.80 | 0.22 | 0.11 | 0.33 |
| TGFB1 | -0.08 | 0.17 | 0.71 | -0.04 | 0.30 | 0.63 |
| TNF | 0.13 | 0.16 | 0.71 | 0.08 | 0.18 | 0.46 |
| TNIP1 | -0.10 | 0.39 | 0.91 | 0.24 | <0.001 | 0.002 |

Beta coefficients from GEE represent increase in relative gene expression level over time (CSA presentation until arthritis development or 2-years of non-progression). Significant changes over time are marked in orange.

**Supplemental Figure 1.** Modelled course of gene expression of 37 cytokines, chemokines and related receptors in CSA patients that progressed to RA (n=18). Cytokines and chemokines were measured at baseline and at time of RA development and for reasons of clarity presented in two plots. No statistically significant changes were observed during follow-up. CSA, clinical suspect arthralgia; RA, rheumatoid arthritis.

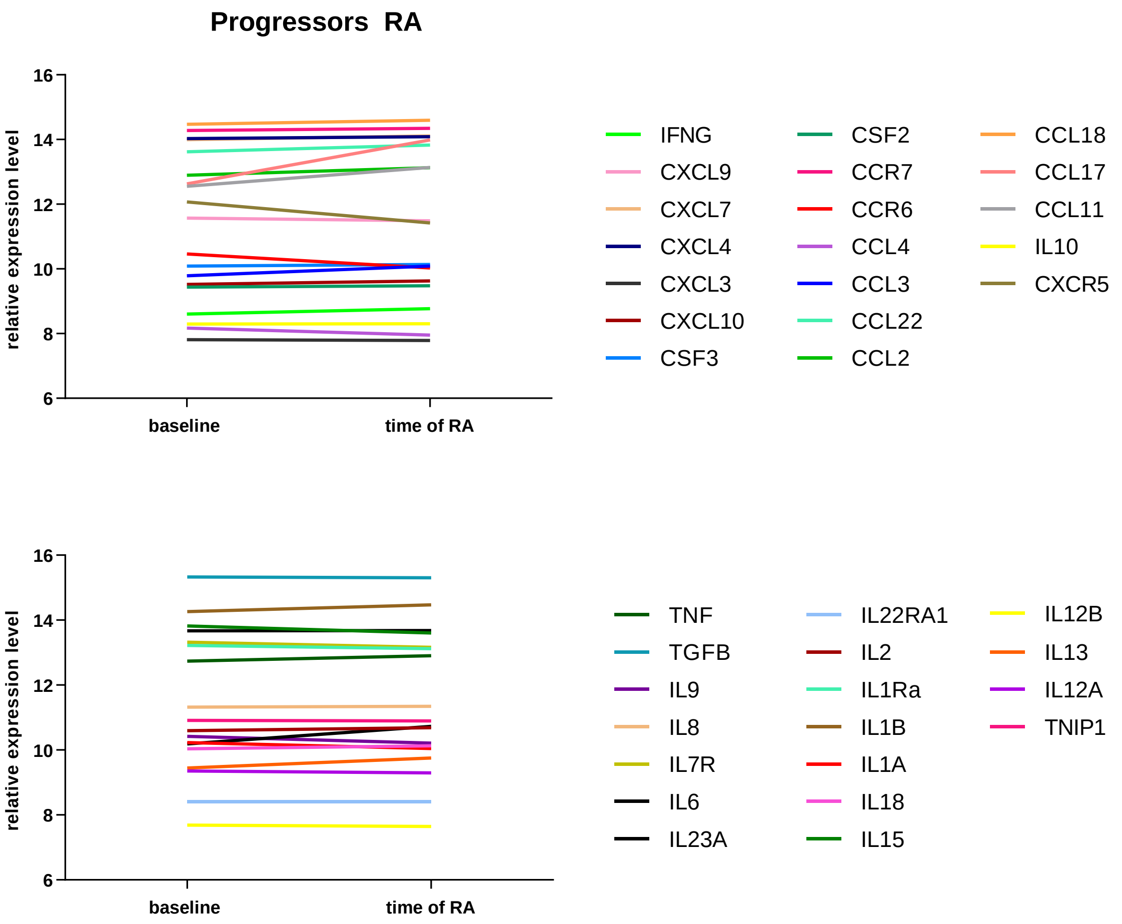

**Supplemental Figure 2.** Modelled course of gene expression of 37 cytokines, chemokines and related receptors in CSA patients that progressed to arthritis in ACPA+ (n=6) and ACPA- (n=16) CSA patients separately. Cytokines, chemokines and related receptors were measured at baseline and at time of arthritis development and for reasons of clarity presented in two plots. No statistically significant changes were observed during follow-up. CSA, clinical suspect arthralgia; IA, inflammatory arthritis.

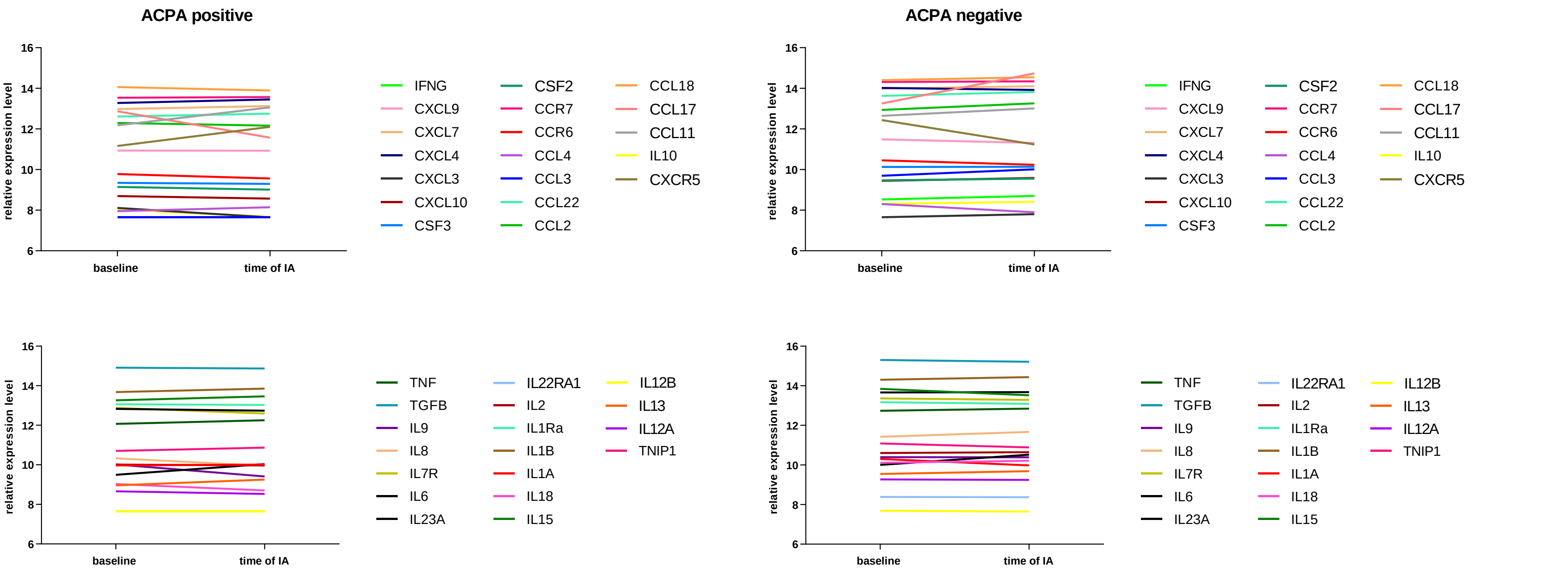

**Supplemental table 3.** Comparison of gene expression at CSA onset between ACPA+ and ACPA- patients that progressed to IA.

| Gene | Beta coefficient | p-value | FDR adjusted p-value |
|---|---|---|---|
| BLR1 | -0.19 | 0.19 | 0.58 |
| CCL11 | -0.09 | 0.57 | 0.83 |
| CCL17 | 0.00 | 0.95 | 0.98 |
| CCL18 | 0.00 | 0.99 | 0.99 |
| CCL2 | -0.20 | 0.33 | 0.61 |
| CCL22 | -0.17 | 0.75 | 0.89 |
| CCL3 | -0.02 | 0.92 | 0.97 |
| CCL4 | -0.49 | 0.14 | 0.54 |
| CCR6 | -0.27 | 0.28 | 0.58 |
| CCR7 | -1.58 | 0.03 | 0.50 |
| CSF2 | -0.85 | 0.14 | 0.54 |
| CSF3 | -1.48 | 0.06 | 0.54 |
| CXCL10 | -0.20 | 0.29 | 0.58 |
| CXCL3L1 | 0.39 | 0.37 | 0.61 |
| CXCL4 | -0.38 | 0.24 | 0.58 |
| CXCL7 | -0.46 | 0.13 | 0.54 |
| CXCL9 | -0.27 | 0.28 | 0.58 |
| IFNG | -0.13 | 0.40 | 0.63 |
| IL10 | 0.22 | 0.37 | 0.61 |
| IL12A | -0.60 | 0.27 | 0.58 |
| IL12B | -0.29 | 0.90 | 0.97 |
| IL13_5RA | -0.96 | 0.01 | 0.36 |
| IL13_IFN | -0.78 | 0.11 | 0.54 |
| IL15 | -0.91 | 0.10 | 0.54 |
| IL18 | -0.06 | 0.69 | 0.88 |
| IL1A | 0.02 | 0.86 | 0.97 |

| | | | |
|---|---|---|---|
| IL1B | -0.43 | 0.46 | 0.70 |
| IL1Ra | -0.24 | 0.65 | 0.87 |
| IL2 | -0.53 | 0.14 | 0.54 |
| IL22RA1 | 0.11 | 0.89 | 0.97 |
| IL23A | -1.64 | 0.06 | 0.54 |
| IL6_5RA | 0.11 | 0.59 | 0.84 |
| IL7R | -0.27 | 0.34 | 0.61 |
| IL8 | -0.09 | 0.66 | 0.87 |
| IL9 | -0.51 | 0.20 | 0.58 |
| TGFB1 | -0.26 | 0.74 | 0.89 |
| TNF | -0.96 | 0.16 | 0.56 |
| TNIP1 | -0.55 | 0.22 | 0.58 |

Beta coefficients are derived from logistic regression models with ACPA-negative groups as a reference. (Borderline) significant genes in uncorrected analyses are marked in orange. None of the genes were significantly different between ACPA+ and ACPA- progressors after correction for multiple testing.
